# Estimated impacts of mandatory sodium reformulation targets on sodium consumption, health outcomes, health sector costs and health equity in Aotearoa New Zealand

**DOI:** 10.64898/2026.11.03.26367203

**Authors:** Kava Fuavao, Christine Cleghorn, Briar L McKenzie, Nisa Widyastuti, Rachael M McLean, Boyd Swinburn, Cliona Ni Mhurchu, Vili Nosa, Helen Eyles

## Abstract

**Aims:** Cardiovascular disease (CVD) is the leading cause of death in Aotearoa New Zealand (NZ), including for Māori, the Indigenous population. Sodium intake is a modifiable CVD risk factor. In 2021, the World Health Organization (WHO) released sodium reduction benchmarks for 58 packaged food categories. We aimed to estimate the health, health-sector cost, and equity impacts of mandating these benchmarks in NZ.

**Methods:** Nationally representative data on household food purchases, packaged food sodium content, and dietary intake were used to estimate baseline sodium consumption (2011) and changes following implementation of the WHO benchmarks. A dietary multi-state life-table model estimated impacts on health-adjusted life years (HALYs) and health system costs (2011 NZ$, 3% discount rate) over the remaining lifetime of the 2011 NZ population (n = 4.4 million). Analyses examined impacts by sex and for Māori.

**Results:** Mandatory reformulation was estimated to reduce sodium intake by 470 mg/day per person (1.18 g salt), a 22% reduction from baseline. Across population groups, age-standardised reductions ranged from 380 to 660 mg/day (22-23%), generating an estimated 55,600 HALYs (95% uncertainty interval 19,100-90,000) and NZ$535.2 million in health-system cost savings (95% UI: 183.0-914.6 million) over the population lifetime. Māori gained 11,600 HALYs (95% UI: 4,900-17,700), equivalent to 17.2 HALYs per 1,000 population, compared with 11.8 per 1,000 among non-Māori.

**Conclusion:** Mandating WHO sodium benchmarks in NZ could substantially reduce sodium intake, improve population health, reduce health sector costs, and advance health equity. Findings support adoption of this strategy in Aotearoa NZ.

**Lay summary:** Requiring food manufacturers in Aotearoa New Zealand to meet World Health Organization sodium (salt) targets could reduce salt intake, prevent disease, save health sector costs, and improve health equity for Māori.

- Mandatory sodium reformulation of packaged foods was estimated to reduce average sodium intake by 470 mg per person per day (equivalent to 1.18 g of salt), a 22% reduction from current levels, resulting in health benefits across the population.
- The policy was estimated to generate 55,600 health-adjusted life years and save NZ$535 million in healthcare costs over the population’s lifetime, with greater health gains per person for Māori than non-Māori, helping reduce health inequities.

## Introduction

Sodium intake demonstrates a dose-response association with blood pressure and is a well-established, modifiable risk factor for hypertension and vascular disease (1–3), with evidence also linking higher intake to gastric cancer and chronic kidney disease (4, 5). Cardiovascular disease (CVD) is the leading cause of death in NZ, and high systolic blood pressure is the largest contributor to CVD burden, accounting for approximately one-quarter to one-third of total CVD disability-adjusted life years and an estimated 5,400 deaths in 2019 (6). The economic cost associated with CVD is substantial, in 2020 approximately $600 million NZD in direct healthcare system costs were attributed to CVD (7).

Hypertension is treatable, however, of an estimated 968,000 NZ adults (30-79 years) with hypertension in 2019 only 63% had been given a formal hypertension diagnosis, 50% had received treatment, and 27% had their hypertension controlled (6, 8). Driven by the impacts of colonisation, Māori (the indigenous population (tangata whenua) of NZ) are inequitably burdened by high blood pressure and related conditions (9, 10). Unfair differences in access to healthy food and the health system have also impacted people of Pacific and South Asian ethnicity living in NZ, who are also disproportionately burdened by CVD (9, 10). These inequities highlight the importance of population-level prevention strategies that reduce exposure to upstream risk factors (9, 11). Effective population blood pressure-lowering interventions have considerable potential to reduce CVD inequities in NZ.

In 2013 all World Health Organization (WHO) member states, including NZ, agreed to a goal of a 30% reduction in population sodium intake by 2025 (12). While some countries have made meaningful progress (12, 13), there is no evidence that any member country has met this target, with the goal subsequently extended to 2030. In NZ, gold standard 24-hour urine collection surveys conducted in 2012 indicated that adults consume around 3400 mg of sodium per day (mg/d), well above the WHO recommended amount of 2000 mg/d or less, with processed and packaged foods being the major dietary source (14, 15). Reducing sodium content in packaged foods is considered an effective and cost effective approach to combat non-communicable diseases (12, 16). In NZ there are limited population level sodium reduction initiatives, and no comprehensive and co-ordinated sodium reduction strategy. While there are two key initiatives that are relevant to sodium, the Health Star Rating front of package labelling scheme (17) and the NZ Heart Foundation food reformulation programme (HeartSAFE) (18), both are voluntary, and have not been applied comprehensively across the packaged food supply, limiting their potential impact (19–21).

To support international efforts to reduce population sodium intake, the WHO has set global benchmarks for sodium levels across numerous packaged food categories (22). First released in 2021 and subsequently updated in 2024, the benchmarks include sodium targets for a wide variety of packaged foods that are key dietary contributors to sodium intake (11 food categories and 58 subcategories). The targets for each category are based on the lowest targets already implemented by individual countries (22). While the impact of full compliance to the NZ HeartSAFE scheme on mean sodium intake has been estimated (260 mg/day reduction) (19), the WHO benchmarks cover a wider range of products (18, 22) and as such, the hypothesis is that application of the WHO benchmarks would have a greater impact on sodium intake. Further, it is not known what impact application of the WHO benchmarks would have on health and health related cost savings in NZ, due to reductions in sodium consumption. The aim of this study was to apply these benchmarks to the NZ packaged food supply to estimate the sodium consumption, health, health equity and health system cost impacts of mandating the WHO sodium benchmarks in NZ.

## Methods

The current study modelled the potential impact of full implementation of the WHO sodium benchmarks on sodium consumption, health adjusted life years gained (HALYs), health equity and health system costs in NZ, following the process below (**Figure 1**).

**Figure 1.**
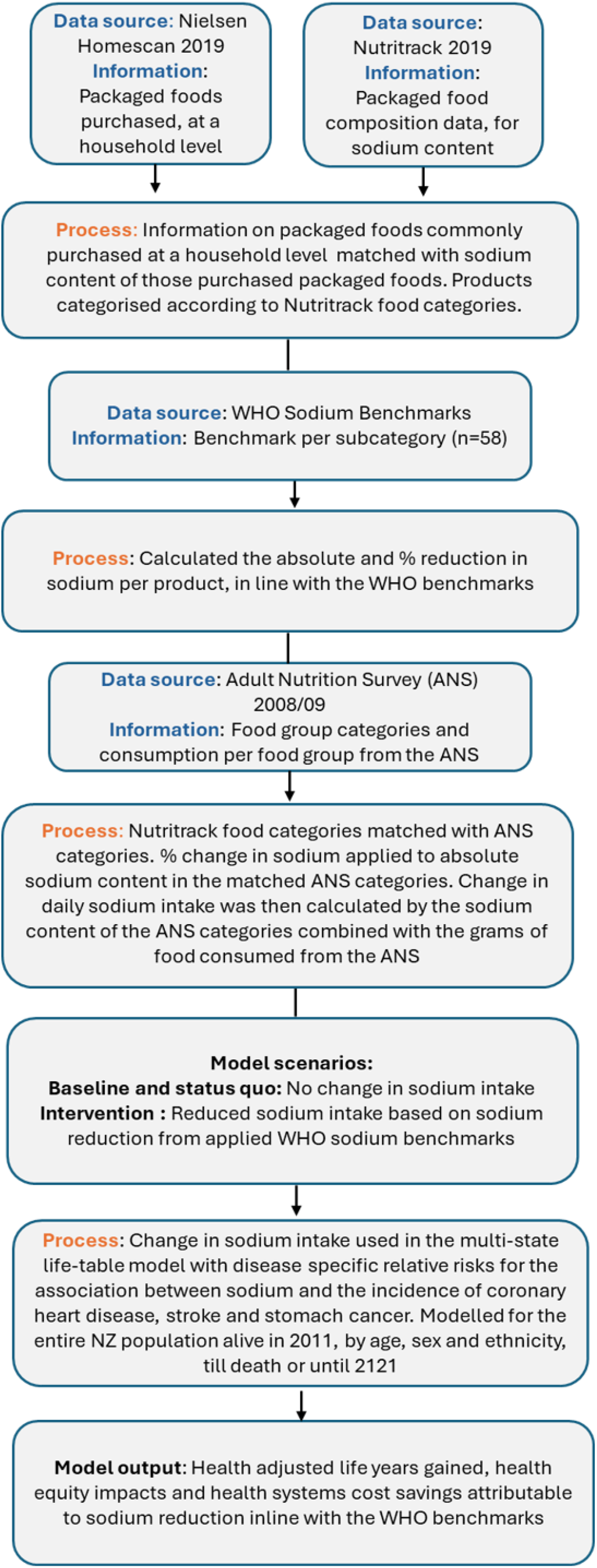
Flow chart of approach to applying the WHO sodium reduction benchmarks to the New Zealand food supply and estimating the related impact on sodium intake, health, equity and health systems cost savings outcomes

### Information used to estimate the impact of the WHO sodium benchmarks on sodium consumption

#### Purchased products, New Zealand Nielsen Homescan

The 2019 New Zealand Nielsen Homescan data was used to identify foods purchased at a household level. The data comprises purchasing information voluntarily submitted by participating households, which is then checked through a standardised procedure to ensure accurate data.

The panel households scan barcodes of consumer goods purchased and brought into the home, encompassing groceries from diverse retail channels like supermarkets, convenience stores, specialist stores, pharmacies, liquor outlets, and online platforms (23). In 2019, 1,800 households from across NZ participated, and 30,900 unique food and beverage products were scanned. From this, 17,527 unique items were excluded for this analysis, based on the following criteria:

- Fresh food products (containing limited or no sodium), n=2,656

- Alcohol products, n=3,702

- Non-food products, n=29

- Products that were <2.5^th^ percentile for sales, n=11,140

This resulted in 13,373 unique packaged foods from 1,799 households (**Supplementary Figure**).

#### Sodium content of purchased products, Nutritrack

Nielsen Homescan data does not contain nutrient content information. As such, food items from Nielsen Homescan were matched with data from 2019 Nutritrack (24). Nutritrack is a database of packaged foods and beverages sold at major supermarkets in New Zealand, capturing nutrition information recorded on the mandatory back of pack product Nutrition Information Panel (NIP). Nutritrack data can be linked to Nielsen data by product barcode. In 2019 17,796 unique packaged food and beverage products were identified. From this, 851 products were excluded based on the following:

• Identical products with different package sizes, n=3

• Products with a NIP reporting reconstituted nutrition information only, n=692

• Products with a problem NIP (for example, products containing infeasible nutrient values (sodium content being extraordinarily high beyond what is physically possible or safe for consumption), n=45

• Products with multiple NIPs (for example, a meal kit with a separate NIP for each component), n=111

This resulted in 16,945 unique packaged food and beverage items (**Supplementary Figure**).

Linking Nielsen Homescan data with Nutritrack data

A total of 9,687 products were matched by barcode between Nielsen and Nutritrack databases. There were 3,686 products from the Nielsen database that did not match to a barcode in Nutritrack. Of these, 3,580 products were manually matched to Nutritrack food categories, with sample means for the missing nutrients calculated based on these categories. One hundred and six products were unable to be matched and were excluded. This resulted in a combined data set of 13,267 packaged food and beverage products, from 1,700 households, providing information on sales weighted sodium purchased.

#### Calculating change in sodium content, based on application of the WHO Sodium Benchmarks

The WHO sodium benchmarks provide target levels for the sodium content of specific food categories, with those categories to either meet the benchmark, or ideally, be below the benchmark. For this study we considered application of the WHO sodium benchmarks as a mandatory policy (i.e., all foods with a relevant sodium benchmark would reduce their sodium content to meet the benchmark if they did not already).

We matched Nutritrack foods with the food categories and subcategories covered in the WHO sodium benchmarks, **Figure 1**.

Baseline:

1. Each product’s total amount of sodium was calculated by dividing its sodium content (mg/100g) by 100 and multiplying it by its packaging size (g)

2. The total amount of sodium purchased was calculated by multiplying each product’s total amount of sodium by the number of purchases for that product.

3. The amount of sodium purchased was summed for all products within each food group

Intervention:

1. Reductions in sodium content, in terms of absolute (mg) and then as a percentage were calculated after intervention, that is all foods identified as a food of interest by the WHO (and with a sodium content above the WHO benchmark) had their average sodium content level reduced, mimicking reformulation, to meet the target. This was conducted for each of the 58 WHO subcategories.

2. The amount of sodium purchased was summed for all products by food group as described above for the new “intervention” total sodium.

Percentage change in sodium content was calculated as the difference between baseline and intervention sodium content for each food group.

#### Converting to sodium consumption using the Adult Nutrition Survey

The most recent New Zealand Adult Nutrition Survey (ANS) was carried out from October 2008 to October 2009, collecting nutrition intake information by 24-hour diet recall from 4,721 New Zealand adults (aged 15 years and older) (25). To convert the household level sales weighted sodium data into sodium consumption, the Nutritrack food categories were matched to the ANS food categories with corresponding food quantity consumption data. Percentage change in sodium content was applied to absolute sodium levels in the matched ANS categories. Change in daily sodium intake was then calculated by the sodium content of the ANS categories combined with the grams of foods consumed from the ANS. This allowed estimation of changes in sodium consumption at an individual level, used to estimate the HALYs gained, equity impacts, and related health system savings.

### Effect of changes in sodium consumption on health adjusted life years and health systems savings

We used a DIET multi-state life-table (MSLT) model to estimate the difference in health adjusted life years (HALYs), health equity impacts and health system costs between the current NZ diet and the same diet where the WHO sodium benchmarks had been applied as a mandatory policy. Details of this model have been described in depth elsewhere (26), the specific approach for the present study is detailed as follows. The total change in daily sodium intake for Māori males, Māori females, non-Māori males and non-Māori females was calculated for use in the MSLT model. The model was stratified into Māori and non-Māori to assess policy impacts for the Māori population and assess the policy’s ability to reduce health inequities and meet Te Tiriti o Waitangi obligations. At the time of model development population numbers for Pacific people were too low to estimate disease rates accurately for the Pacific population. The change in sodium intake was combined with disease specific relative risks sourced from the Global Burden of Disease study, for the association between sodium and incidence of coronary heart disease (CHD), stroke and stomach cancer. The proportion of the population (by sex, age, and ethnicity) in each disease life table is a function of the disease incidence and case fatality, with remission rates also considered for cancers (21).

In the model the entire NZ population, alive in 2011 (N = 4.4 million), classified by sex, age and ethnicity (Māori and non-Māori) was modelled out to death or until year 2121. The intervention was modelled as if it were put in place in the base year (2011) and kept in place indefinitely. The ‘base case’ model used 3% discounting and took a health system costing perspective. All disease input parameters were specified by sex, age and ethnicity where data were available. Input parameters have been previously published (27).

Individually-linked data from publicly-funded (and some privately-funded) health events occurring in 2006–10 were used to calculate age and sex specific health system costs in NZ$ for 2011 (26). These costs included hospitalisations, inpatient procedures, outpatients, pharmaceuticals, laboratories and expected primary care usage. Costs that were assigned in the model fell into the following three categories: (i) Sex and age-specific annual cost of a citizen who does not have a sodium-related disease and is not in the last six months of their life; (ii) Disease-specific excess costs for people in the first year of diagnosis, last six months of life if dying of the given disease, and otherwise prevalent cases of each disease in the model; (iii) Costs associated with the last six months of life if dying from a disease not in the model (26).

Microsoft Excel using an Ersatz add-in (Epigear International, created by Jan J Barendregt (28)) was used to run the policy with uncertainty. Each of the 2000 simulations involved a random draw from the probability density function about those parameters specified with uncertainty (27). The main results produced were incremental HALYs gained and net health system cost savings. The health system cost savings reflect projected future health system savings resulting from decreases in disease incidence due to the WHO sodium benchmarking policy, including extra health costs from any increased life span.

### Sensitivity Analysis

The impact of colonisation on the Māori population resulted in a higher morbidity and mortality rate than the non-Māori population, thus diluting the potential health gains and creating a disadvantage in the main analyses where impacts for Māori could appear less than they would if Māori had the same life expectancy as non-Māori. As such, an equity analysis was conducted, equally valuing health gain in Māori to that of non-Māori (29). For the equity analysis the underlying morbidity and mortality rates for Māori were set to non-Māori values to estimate impacts as if these inequities were not present.

## Results

### Estimated impact of WHO benchmarks on sodium content of foods

In the intervention scenario, application of the WHO benchmarks resulted in sodium reductions of 40% or greater for the food categories: “pies and pastries” (66% reduction), “sausages and processed meats” (52%), “puddings and desserts” (47%), and “beef and veal” (44%). The smallest reductions at a food category level were seen for “bread-based dishes” (5%) and “potatoes, kumara and taro” (8%), **Table 1**.

**Table 1.**
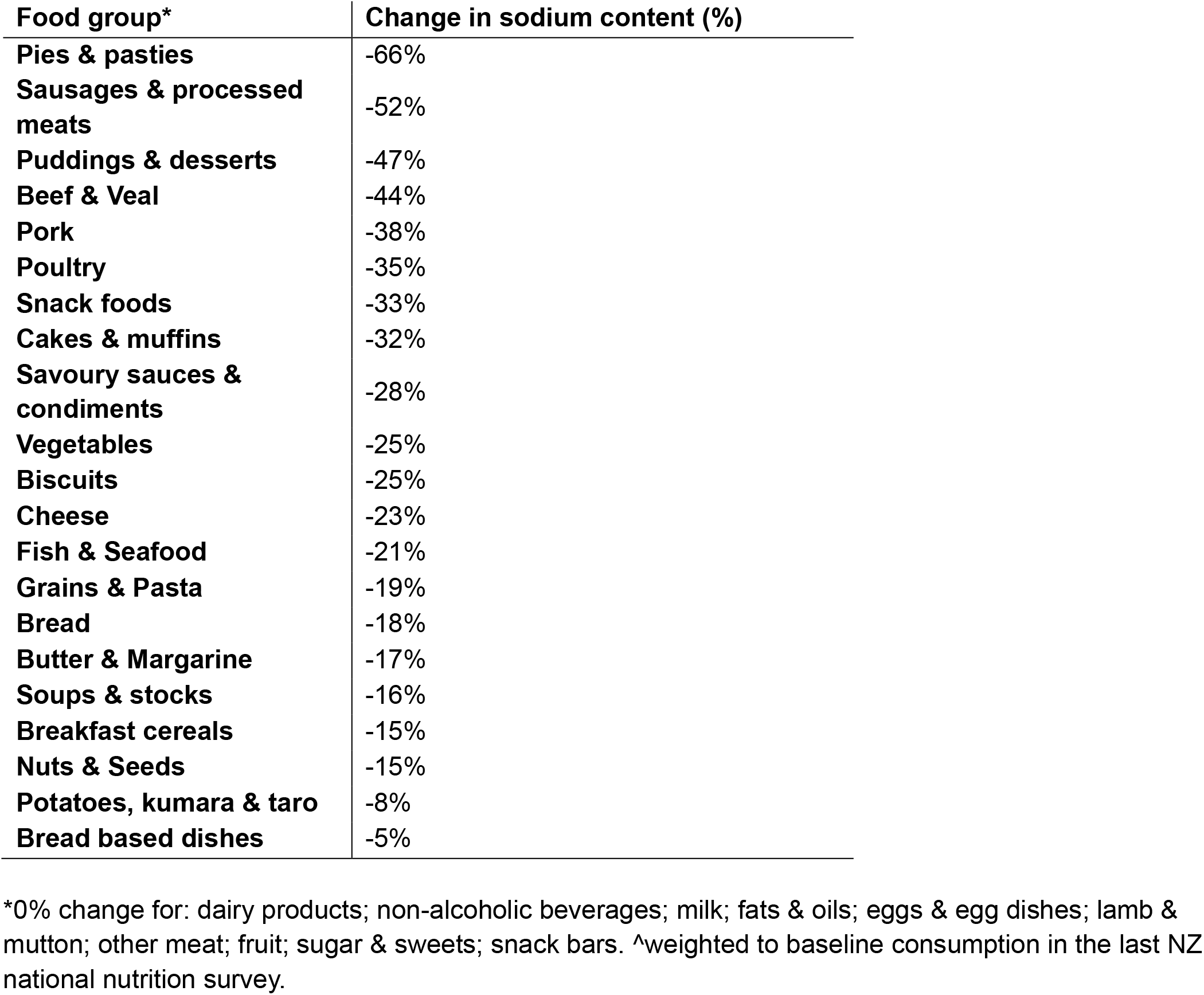
Percent change in sodium content by food group.

### Estimated impact of WHO benchmarks on sodium intake

With application of the benchmarks, an estimated average reduction in sodium intake of 470 milligrams per person per day (mg/day), equivalent to 1.18 g of salt, could be achieved, corresponding to a 22% reduction from baseline (2011) intake levels, **Table 2**. For Māori, there was an estimated 660 mg/day reduction in sodium intake for males, and a 410 mg/day reduction for females. For both sexes, this corresponds to a 23% reduction in intake from baseline levels. For non-Māori, males had an estimated 530 mg/day reduction in sodium intake, and females had an estimated 380 mg/day reduction, both corresponding to a 22% reduction from baseline.

**Table 2.** Estimated reduction in sodium intake, following application of the WHO sodium benchmarks.

|  | Overall | Māori |  | Non- Māori |  |
|---|---|---|---|---|---|
|  |  | Males | Females | Male | Females |
| Reduction in sodium intake (mg/day) | 470 | 660 | 410 | 530 | 380 |
| Equivalent salt reduction (g/day) | 1.18 | 1.65 | 1.03 | 1.33 | 0.95 |
| Percent reduction from baseline (%) | 22 | 23 | 23 | 22 | 22 |

### Estimated impact of reformulation on health gains and health system savings

The estimated reductions in sodium intake were associated with moderate gains in HALYs across the total NZ population, over the life course of those alive in 2011, at 55,600 (95% Uncertainty Interval (UI): 19,100 to 90,000) and substantial predicted health system cost savings of $535.2 Million(M) (95% UI: 183.0 to 914.6) over the same time period, **Table 3**. Overall, HALYs gained were higher for males (35,400) than for females (20,200). For Māori, 11,600 (4,900 to 17,700) HALYs could be gained, higher for males in comparison to females (7,000 (3,400 to 10,100) vs 4,600 (1,500 to 7,700)). For non-Māori, 44,000 (14,300 to 72,800) HALYs could be gained, with 28,400 (10,700 to 44,800) HALYs gained for males and 15,600 (3,600 to 28,600) HALYs gained for females. Per capita, these findings correspond to 17.2 HALYs gained per 1,000 people for Māori and 11.8 HALYs gained per 1,000 people for non-Māori.

**Table 3.** Estimated impact of the WHO sodium reduction benchmarks on daily sodium intake, health adjusted life years (HALYs) and health systems costs.

|  | HALYs Gained |  |  |  | Net health system cost savings (NZ\$ million) |
|---|---|---|---|---|---|
| Population | Total | Māori | Equity analysis for Māori | Non-Māori | Total |
| Total | 55,600 (19,100, 90,000) | 11,600 (4,900, 17,700) | 16,400 (6,500, 25,500) | 44,000 (14,300, 72,800) | 535.2 (183.0 to 914.6) |
| Males | 35,400 (14,000 to 55,000) | 7,000 (3,400 to 10,100) | 9,900 (4,500 to 14,500) | 28,400 (10,700 to 44,800) | 359.9 (134.1 to 593.1) |
| Females | 20,200 (5,100 to 36,000) | 4,600 (1,500 to 7,700) | 6,500 (2,000 to 11,200) | 15,600 (3,600 to 28,600) | 175.4 (45.5 to 323.1) |
| *Per capita* | 12.6 | 17.2 (22.5) | 24.3 (31.9) | 11.8 (14.0) | 121.50 |

All gains are for the 2011 population over their life course, 3% discounting, generated from changes in CHD, stroke, and stomach cancer incidence. \*95% uncertainty intervals in brackets. \*\*per capita results: HALYs/1000 people & $/person. Crude HALYs with age standardised HALYs in brackets.

Most of the health gain seen in this policy came from the prevention of CHD (30,084 HALYs (56% of total HALYs)), then stroke (18,854 HALYs (36%)), with health gain attributed to prevention of stomach cancer as the smallest proportion (4,273 HALYs (8%)), **Figure 2**.

**Figure 2.**
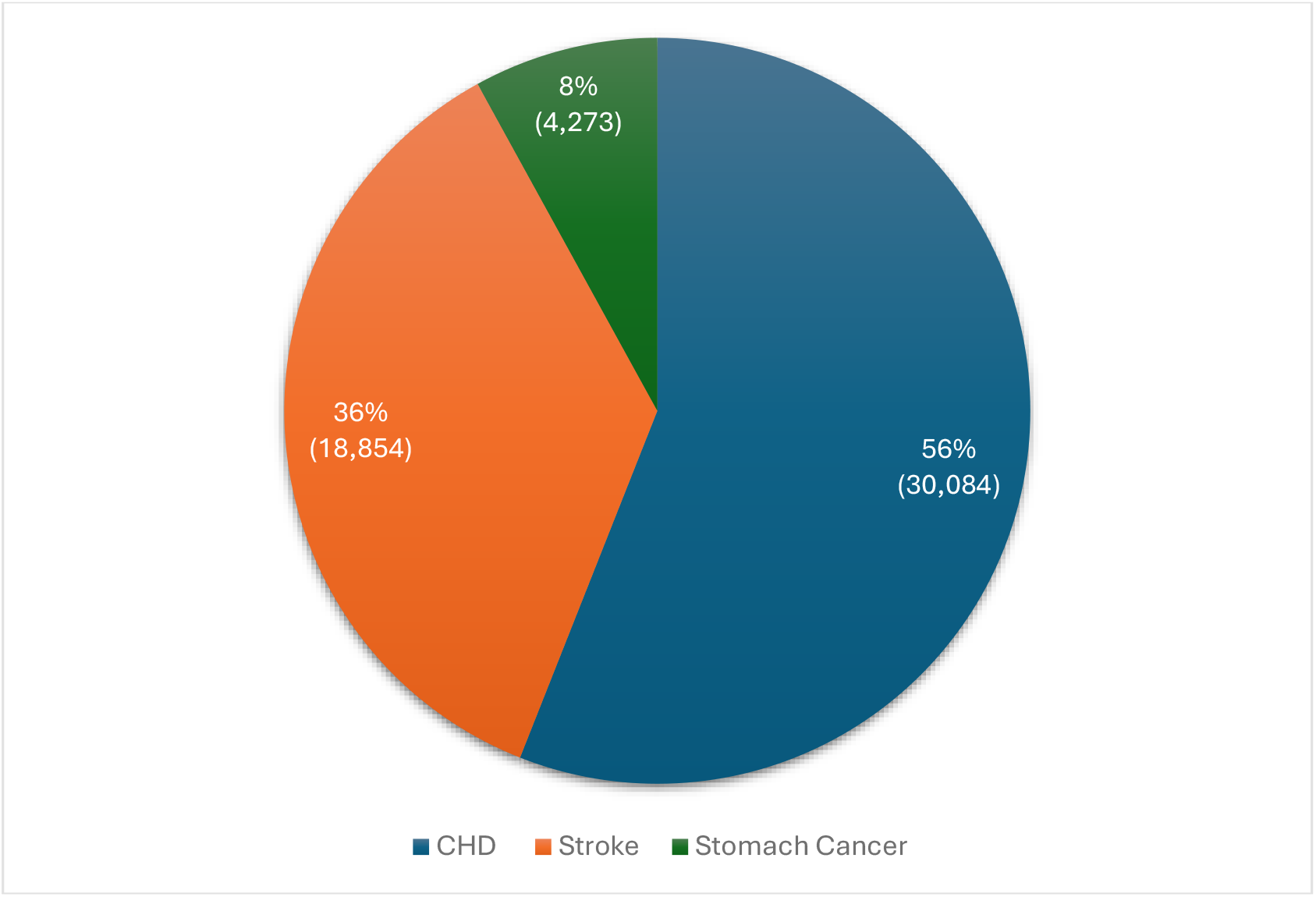
Proportion of health adjusted life years (HALYs) gained attributable to specific diseases on application of the WHO sodium benchmarks (% (HALYs))

### Sensitivity analysis

In the equity analysis, where the underlying morbidity and mortality rates for Māori were set to non-Māori values, an estimated 16,400 (6,500 to 25,500) HALYs were gained for Māori, with 9,900 (4,500 to 14,500) HALYs gained for males and 6,500 (2,000 to 11,200) for females. Overall, corresponding to 24.3 HALYs gained per 1000 people, **Table 3**.

## Discussion

Our modelled scenario, in which the WHO sodium benchmarks are implemented as a mandatory food reformulation policy in New Zealand, resulted in a meaningful reduction in sodium intake of 470 mg/day per person (a 22% reduction). This translated into modest but important reductions in disease burden, with an estimated 30,084 health-adjusted life years (HALYs) gained from preventing coronary heart disease, 18,854 HALYs from stroke, and 4,273 HALYs from stomach cancer. An estimated 17.2 HALYs per 1000 people were gained for Māori (*24.3 HALYs per 1000 people in the equity analysis*), and 11.8 HALYs gained per 1000 people for non-Māori, indicating direct benefit to the Māori population. The intervention generated substantial health system cost savings, estimated at $535.2 million over the population’s lifetime. Together, these findings highlight the potential of population level sodium reformulation as an effective strategy for reducing the burden of diet-related disease in NZ.

Given the substantial burden of sodium-related disease experienced by Māori in NZ (9, 10), it was important to assess the potential impacts of the intervention for Māori specifically. Twenty-one percent of the total health gain accrued to Māori (37% in the equity analysis), despite Māori comprising approximately 15% of the NZ population in the 2013 NZ census, the closest census conducted to the model base year of 2011 (30). On a per-person basis, for Māori the intervention generated 0.017 HALYs in the main analysis and 0.024 HALYs per person in the equity analysis. These findings are consistent with previous NZ modelling demonstrating that sodium reduction interventions may contribute to reducing health inequities. Nghiem et al. (31) similarly reported modelled health gains for Māori across a range of sodium reduction interventions. In that study, the largest gains for Māori were estimated for the full replacement of regular salt in the packaged food supply with reduced-sodium salt substitutes, resulting in 0.163 QALYs gained per person, while the introduction of strict sodium limits for bread (280 mg sodium per 100 g) generated approximately 0.024 QALYs per person, greater than findings in the present study. Taken together, these findings suggest that policies that reduce the sodium content of foods have the potential to contribute to health gains for Māori in NZ.

This is the first study to estimate the potential effects of applying the WHO sodium benchmarks on health outcomes and health system costs in NZ. A previous NZ study examined the potential impacts of full compliance with the HeartSAFE Heart Foundation reformulation targets (19). That study assessed effects on sodium intake only and estimated that full compliance could reduce intake by 260 mg/day, which is smaller than the 470 mg/day reduction estimated in the present study. This difference is likely due to the HeartSAFE targets applying to fewer food categories and in some cases having less stringent targets than the WHO benchmarks, making it plausible that the HeartSAFE targets would generate proportionally smaller health gains than those estimated here. Our findings are broadly consistent with modelling studies from other countries, which similarly suggest that sodium reformulation could deliver meaningful health and economic benefits. In Australia, full implementation of the WHO benchmarks was estimated to reduce mean sodium intake by 404 mg/day and prevent mortality and morbidity, including approximately 1,770 deaths annually (32). In Canada, modelling of a scenario in which sodium intake was reduced to 2,000 mg/day (the WHO recommended limit) estimated that 3,252 cardiovascular disease deaths (95% UI 1,380–5,321) could be averted, corresponding to 5.6% of all CVD deaths in Canada in 2019 (33). Similarly, modelling of the UK’s country-specific sodium targets, which apply to a wider range of products than the WHO benchmarks by including foods sold in restaurants and fast-food outlets (34), estimated that mandatory implementation could reduce sodium intake by 17.5% and generate net savings of more than £1 billion for the National Health Service (35). Although the magnitude of predicted benefit varies by country context and the specific reformulation scenario modelled, the overall direction of evidence is consistent. Taken together, these findings reinforce that mandatory, population level sodium reformulation targets are likely to be beneficial from both a health and an economic perspective.

Modelling studies produce important estimates of the likely impact of different policy scenarios; however, real world evaluations are needed to show whether sodium targets can be implemented at scale, and how this translates into reductions in population intake. In 2019, 19 countries had mandatory maximum sodium limits for foods (13), but few policies covered a broad range of packaged food products, and even fewer have been comprehensively evaluated. South Africa provides one example of mandatory sodium regulation in practice. The government introduced sodium regulations in 2013, applying to 13 categories of processed foods, using a stepwise implementation approach to support the food industry in meeting final sodium targets (36).

Monitoring of the food supply showed that, in 2021, 75% of products had sodium levels at or below the final targeted limits (36). Although not designed as a formal policy evaluation, a survey using 24-hour urine samples found a 460 mg/day reduction in sodium intake among adults with a median age of 54 years between 2015 and 2019 (37). These findings suggest that mandatory reformulation can be translated into meaningful reductions in population sodium intake. The UK salt reduction programme, established in 2003/04, is also widely cited as a successful sodium reduction strategy (38). This was a voluntary programme, but one of its central components was the setting of progressively lower salt targets for different food categories, with clear timeframes for industry compliance. Over a seven-year period, the programme achieved a 15% reduction in sodium intake, as measured by 24-hour urine collections, from 3,800 mg/day to 3,240 mg/day (a 560 mg/day reduction) (38). Despite this reduction, mean population sodium intake in the UK remains well above recommended levels, and efforts to further reduce intake continue (35, 39).

Together, these examples suggest that sodium reformulation targets can reduce sodium intake at the population level, particularly when they are mandatory and are accompanied by strong implementation and monitoring systems. At the same time, they also highlight the need for additional interventions to support sodium reduction, with reformulation being one component of a broader coordinated sodium reduction strategy.

To support Member States in achieving the 2030 sodium reduction targets, WHO developed the SHAKE technical package (16). SHAKE is intended to guide governments in leading comprehensive sodium reduction programmes and emphasises the importance of a government-led approach that limits industry influence. The package outlines seven key strategies (16): food reformulation; front-of-pack labelling; food procurement and food service policies (including limiting the sale or provision of high-sodium foods in public settings and government food programmes); restrictions on food marketing; taxation of unhealthy foods; behaviour change communication and mass media campaigns to promote healthy diets; and the replacement of regular salt with lower-sodium salt substitutes. The INTERSALT study reported that a 100 mmol/day reduction in sodium intake was associated with at least a 2.2 mmHg reduction in systolic blood pressure, a change linked to meaningful reductions in cardiovascular disease mortality (40). Although the sodium reduction estimated in our study was more modest (equivalent to 20.4 mmol/day), it would still be expected to contribute to improved cardiovascular health.

However, achieving larger reductions in blood pressure and disease burden will likely require reformulation to be implemented alongside additional sodium reduction interventions. The previous modelling by Nghiem et al. (31) estimated the impact of a range of sodium reduction initiatives that align with some of the SHAKE recommendations, in the NZ context and considering impacts for Māori. Compared with the scenario modelled here, interventions such as replacing regular sodium salt with potassium-enriched salt across the food supply or applying stricter sodium targets specifically to bread (a key contributor to sodium intake in NZ) generated larger reductions in sodium intake and related health gains than the application of the WHO benchmarks alone, with proportionally higher health gains for Māori (31). Collectively, the findings from this study and from previous NZ modelling illustrate the value of a package of complementary sodium reduction interventions. Consistent with the WHO SHAKE guidelines (16), these results suggest that such an approach would lead to national level health gains and health system cost savings.

### Strengths and limitations

This is the first study to model the health and health sector cost savings resulting from mandatory application of the WHO benchmarks in NZ. Nationally representative data sources have been utilised to inform this modelling, including foods purchased and sodium content, sodium intake, disease burden and health sector related costs, utilising the most relevant model input data. The model used was developed specifically for the NZ context and has been used to estimate the impact of other diet related policies in NZ (21, 31). There are important limitations in our approach to consider in line with our findings. Changes in sodium intake because of reformulation were estimated through use of the ANS 2008/09 (25). This is the most recent national nutrition survey in NZ. However, it is likely that diets have changed since 2008/09. There is evidence of an increased reliance globally on ultra-processed foods (41), with studies conducted in the UK indicating that higher ultra-processed food intake is linked with higher sodium intake (42). This means our findings may underestimate the potential health impacts of reformulation. Further, the WHO benchmarks have since been updated to be stronger (lower sodium limits, across a broader range of food categories) (43), meaning there would be a greater predicted impact from their application compared to the targets applied in the present study. Our equity analysis was conducted for Māori, however, inequities in exposure to diet related disease risk are prevalent for other groups within the NZ population, and broader equity considerations would be important to include in future studies. Our approach also assumed that people do not respond to reformulation by adding more salt to food during cooking or when eating. Evidence suggests that gradual reformulation of products to contain less sodium is often not detected by consumers (44). Finally, implementation costs were not included when modelling the impact on health sector costs. However, if implementation costs were below NZD$530 million, the policy would be considered cost saving and if the implementation costs were below NZD$2.5 billion the policy would be considered cost effective. Economic modelling of application of the WHO benchmarks in the Australian context suggest that implementation costs would be approximately AUD$55 million (approximately NZD$66 million) across the population lifetime, indicating that it would be cost saving and effective in the NZ context (45).

## Conclusion

This study modelled the health and health system cost savings associated with mandating the WHO sodium benchmarks in New Zealand, estimating meaningful reductions in sodium intake and modest but important reductions in the related disease burden. The intervention was equitable, importantly generating more health gains for Māori than non-Māori, with the potential to generate substantial health system cost savings. Overall, these findings demonstrate that sodium reformulation, aligned with food category specific sodium targets, would be an important component of a broader, government-led sodium reduction strategy to support improved population health in Aotearoa New Zealand.

## Data Availability

All data produced in the present study may be sought upon reasonable request to the authors.

## Acknowledgements

We acknowledge and thank the following: Julia Lyon for early collaboration and support. Essa Tawfiq for their support with data linkage. Imogen Roux for their support in applying the WHO benchmarks. The Nutritrack data collectors and project management team. Those who contributed to models used in this work (Tony Blakely, Nick Wilson, Linda Cobiac, Anja Mizdrak, Giorgi Kvizhinadze, Nhung Nghiem).We thank the 4,721 New Zealanders who participated in the 2008/09 New Zealand Adult Nutrition Survey. The New Zealand Ministry of Health funded the 2008/09 New Zealand Adult Nutrition Survey. Collection of Nutritrack data were funded by a Health Research Council of New Zealand Programme Grant to C.N.M (18/672). Access to some of the data used in this study was provided by Statistics New Zealand under conditions designed to keep individual information secure in accordance with requirements of the Statistics Act 1975.

Data interpretation does not necessarily represent an official view of Statistics New Zealand. Artificial Intelligence (AI) was used via Microsoft 365 CoPilot to suggest a layout and animations for the graphical abstract submitted with this manuscript and to provide suggested grammatical changes to aid readability in specific sections of the manuscript discussion only. It was not used outside of these two areas or to interpret findings or generate new information.

---

**Supplementary Figure.**
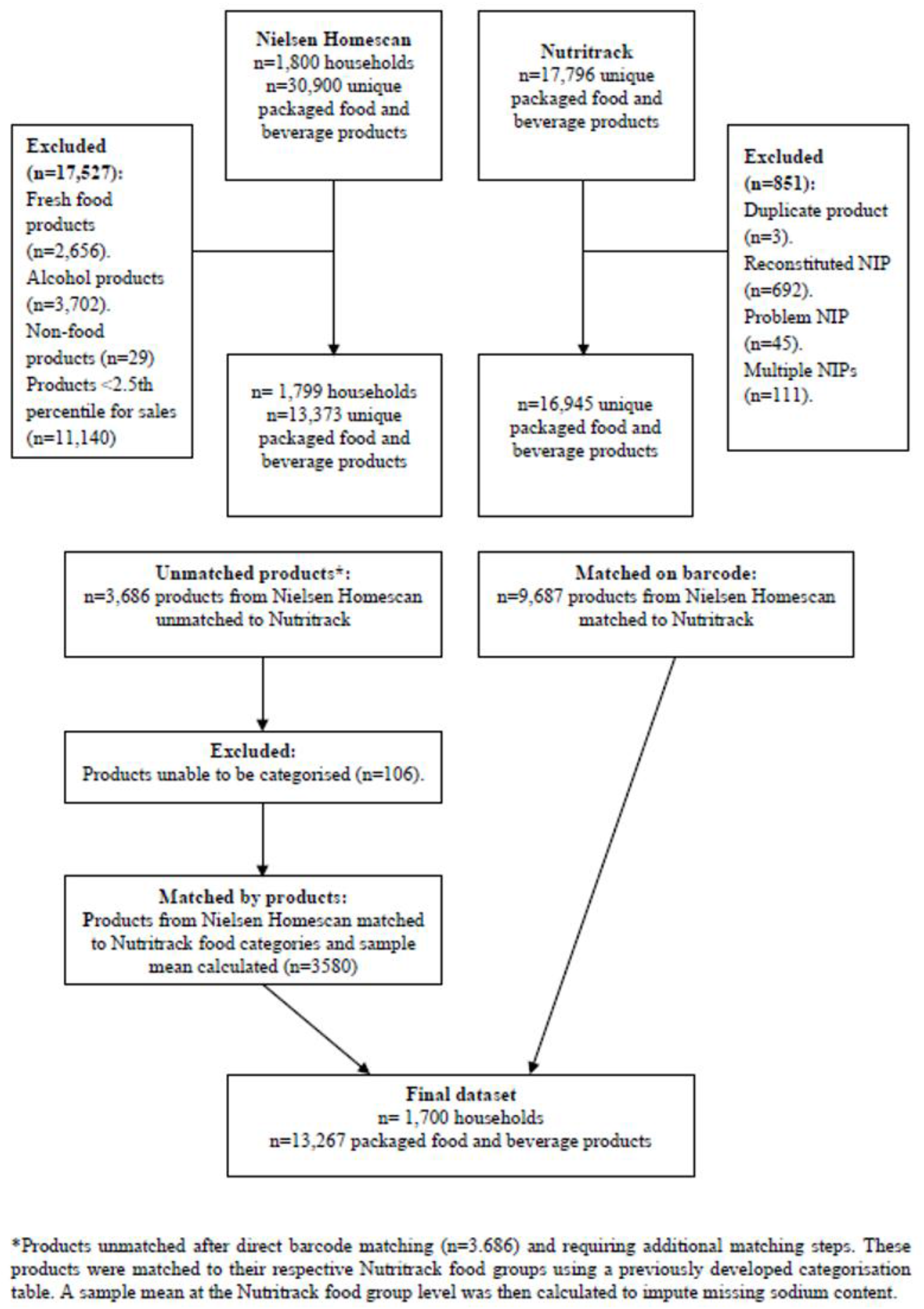
Flow chart showing exclusion criteria and matching results for matching New Zealand Nielsen Homescan data to Nutritrack food composition data

